# Pre-infection neutralizing antibodies, Omicron BA.5 breakthrough infection, and long COVID: a propensity score-matched analysis

**DOI:** 10.1101/2023.04.05.23288162

**Authors:** Shohei Yamamoto, Kouki Matsuda, Kenji Maeda, Kumi Horii, Kaori Okudera, Yusuke Oshiro, Natsumi Inamura, Takashi Nemoto, Junko S. Takeuchi, Yunfei Li, Maki Konishi, Kiyoto Tsuchiya, Hiroyuki Gatanaga, Shinichi Oka, Tetsuya Mizoue, Haruhito Sugiyama, Nobuyoshi Aoyanagi, Hiroaki Mitsuya, Wataru Sugiura, Norio Ohmagari

**Affiliations:** Department of Epidemiology and Prevention, Center for Clinical Sciences, National Center for Global Health and Medicine, Tokyo, Japan; AIDS Clinical Center, National Center for Global Health and Medicine, Tokyo, Japan; Japan Foundation for AIDS Prevention, Tokyo, Japan; Division of Antiviral Therapy, Joint Research Center for Human Retrovirus Infection, Kagoshima University, Kagoshima, Japan; Department of Refractory Viral Infection, Research Institute, National Center for Global Health and Medicine, Tokyo, Japan; Infection Control Office, Center Hospital of the National Center for the Global Health and Medicine, Tokyo, Japan; Infection Control Office, Kohnodai Hospital of the National Center for the Global Health and Medicine, Chiba, Japan; Department of Laboratory Testing, Center Hospital of the National Center for the Global Health and Medicine, Tokyo, Japan; Department of Academic-Industrial Partnerships Promotion, Center for Clinical Sciences, National Center for Global Health and Medicine, Tokyo, Japan; Center Hospital of the National Center for the Global Health and Medicine, Tokyo, Japan; Kohnodai Hospital of the National Center for the Global Health and Medicine, Chiba, Japan; Center for Clinical Sciences, National Center for Global Health and Medicine, Tokyo, Japan; Disease Control and Prevention Center, National Center for Global Health and Medicine, Tokyo, Japan

## Abstract

**Importance:** Investigating the role of pre-infection humoral immunity against Omicron BA.5 infection risk and long COVID development is critical to inform public health guidance.

**Objective:** To investigate the association between pre-infection immunogenicity after the third vaccine dose and the risks of Omicron BA.5 infection and long coronavirus disease.

**Design, Setting, and Participants:** This nested case-control analysis was conducted among tertiary hospital staff in Tokyo, Japan who donated blood samples in June 2022 (1 month before Omicron BA.5 dominant wave onset [July–September 2022]) approximately 6 months after receiving the third dose of the historical monovalent coronavirus disease 2019 mRNA vaccine.

**Exposures:** Live virus-neutralizing antibody titers against Wuhan and Omicron BA.5 (NT_50_) and anti-SARS-CoV-2 spike protein antibody titers with Abbott (AU/mL) and Roche (U/mL) assays at pre-infection.

**Main Outcomes and Measures:** Symptomatic SARS-CoV-2 breakthrough infections during the Omicron BA.5 dominant wave vs. undiagnosed controls matched using a propensity score. Incidence of long COVID (persistent symptoms ≥4 weeks after infection) among breakthrough infection cases.

**Results:** Anti-spike antibody titers were compared between 243 breakthrough infection cases and their matched controls among the 2360 staff members who met the criteria. Neutralizing antibodies in 50 randomly selected matched pairs were measured and compared. Pre-infection anti-spike and neutralizing antibody titers were lower in breakthrough cases than in undiagnosed controls. Neutralizing antibody titers against Wuhan and Omicron BA.5 were 64% (95% CI: 42–77) and 72% (95% CI: 53–83) lower, respectively, in breakthrough cases than in undiagnosed controls. Individuals with previous SARS-CoV-2 infections were more frequent among undiagnosed controls than breakthrough cases (19.3% vs. 4.1%), and their neutralizing antibody titers were higher than those of infection-naïve individuals. Among the breakthrough cases, pre-infection antibody titers were not associated with the incidence of long COVID.

**Conclusions and Relevance:** Pre-infection immunogenicity against SARS-CoV-2 may play a role in protecting against the Omicron BA.5 infection, but not in preventing long COVID.

**Key Points:** 

**Question:** Does pre-infection anti-SARS-CoV-2 humoral immunity protect against Omicron BA.5 infection and long-COVID development?

**Findings:** Pre-infection neutralizing antibody titers against Omicron BA.5 were lower in subsequent Omicron BA.5 breakthrough infection cases than in matched controls in this nested case-control study of healthcare workers who received the third dose of historical COVID-19 mRNA vaccines approximately 6 months prior. Pre-infection antibody titers could not predict the incidence of long COVID among breakthrough infection cases.

**Meaning:** Higher pre-infection humoral immunity approximately 6 months after the third vaccination may correlate with protection against Omicron BA.5 infection but not against long-COVID development.

## Introduction

mRNA vaccines are effective in lowering the risk of severe acute respiratory syndrome coronavirus 2 (SARS-CoV-2) infection;^1^ however, whether a higher level of vaccine-induced pre-infection humoral immunity is associated with a lower risk of SARS-CoV-2 infection remains unclear.^2-4^ In our previous nested case-control studies, neutralizing antibody titers 1–2 months after the second and third doses of vaccines were not associated with the subsequent risk of SARS-CoV-2 infection during the Delta^3^ and Omicron BA.1/BA.2^4^ epidemics, respectively. This lack of association is partly attributed to the similarities among participants in the determinants of post-vaccine infection risk (same vaccine dose and a short period between vaccination and infection).

The largest epidemic wave of Omicron BA.5 occurred between July and September 2022 in Japan, approximately 6 months after the onset of the third vaccination campaign, and recorded the highest weekly number of cases worldwide.^5^ This may be attributed to the high transmissibility of this variant^6, 7^ and the waning of third-dose vaccine immunogenicity over time.^7, 8^ We performed a serosurvey among the staff of the National Center for Global Health and Medicine (NCGM), Tokyo, Japan in June 2022 (1 month before the start of the Omicron BA.5 epidemic) and stored blood samples. This situation prompted us to test the hypothesis that pre-infection humoral immunity, which may vary considerably among participants 6 months after the third vaccination, can predict the risk of Omicron BA.5 infection.

Beyond the role of pre-infection humoral immunity in preventing SARS-CoV-2 infection, its role for the development of long COVID is largely unknown. A low humoral immune response in the acute phase of SARS-CoV-2 infection has been linked to an increased risk of long COVID in non-vaccinated patients.^9, 10^ However, whether pre-infection vaccine-induced humoral immunity is associated with the risk of long COVID is unclear.

Here, we compared the live-virus and pre-infection neutralizing antibody titers among staff members infected with SARS-CoV-2 during the Omicron BA.5 epidemic and their rigorously matched controls in a well-defined cohort of third-vaccine recipients. Additionally, we investigated the association between the pre-infection neutralizing capacity and long COVID.

## Methods

### Study setting

A repeat serological study was conducted at the NCGM in Japan in July 2020 to monitor the spread of SARS-CoV-2 infection among staff during the COVID-19 epidemic. The details of this study have been reported elsewhere.^3, 4^ In summary, we have completed seven surveys as of December 2022, where we measured anti-SARS-CoV-2 nucleocapsid-(all surveys) and spike-protein antibodies (from the second survey onward) for all the participants using both Abbott and Roche assays, stored serum samples at -80□, and collected information on COVID-19–related factors (vaccination, occupational infection risk, infection prevention practices, behavioral factors, etc.) via a questionnaire. We collected information on long COVID from participants with a history of COVID-19 via the seventh questionnaire survey conducted in December 2022. The self-reported vaccination status was validated using objective information provided by the NCGM Labor Office. We identified COVID-19 cases among the study participants from the COVID-19 patient records documented by the NCGM Hospital Infection Prevention and Control Unit, which provided information on the date of diagnosis, diagnostic procedures, possible route of infection (close contact), symptoms, hospitalizations, return to work for all cases, and virus strain and cycle threshold (Ct) values for those diagnosed at the NCGM. Written informed consent was obtained from all the participants. This study was approved by the NCGM Ethics Committee (approval number: NCGM-G-003598).

### Case-Control Selection

We conducted a case-control study among the staff who participated in the sixth survey conducted in June 2022 and completed three doses of the monovalent mRNA COVID-19 vaccine (any dose pattern of BNT162b2 or mRNA-1273) (**Figure 1**). Of the 2,727 participants, 2,360 received three doses of the mRNA COVID-19 vaccines and donated blood samples. Of those, we identified 262 (11%) breakthrough infection cases, defined as those diagnosed at least 14 days after the third dose by September 21, 2022, using the in-house COVID-19 registry. We selected 243 patients with symptomatic SARS-CoV-2 infections as cases for analysis, after excluding 19 patients with asymptomatic infections. Finally, 243 cases and 2,098 undiagnosed participants formed the basis for the case-control study. We selected a control for each case using propensity score matching, and 243 matched pairs were selected and included in the analysis to compare pre-infection anti-spike antibody titers between cases and controls. The details of the case-control matching algorithm are described in **eText 1**. Of the 243 matched pairs, we randomly selected 50 pairs and measured live virus–neutralizing antibody titers to compare neutralizing antibodies between the groups.

**Figure 1.**
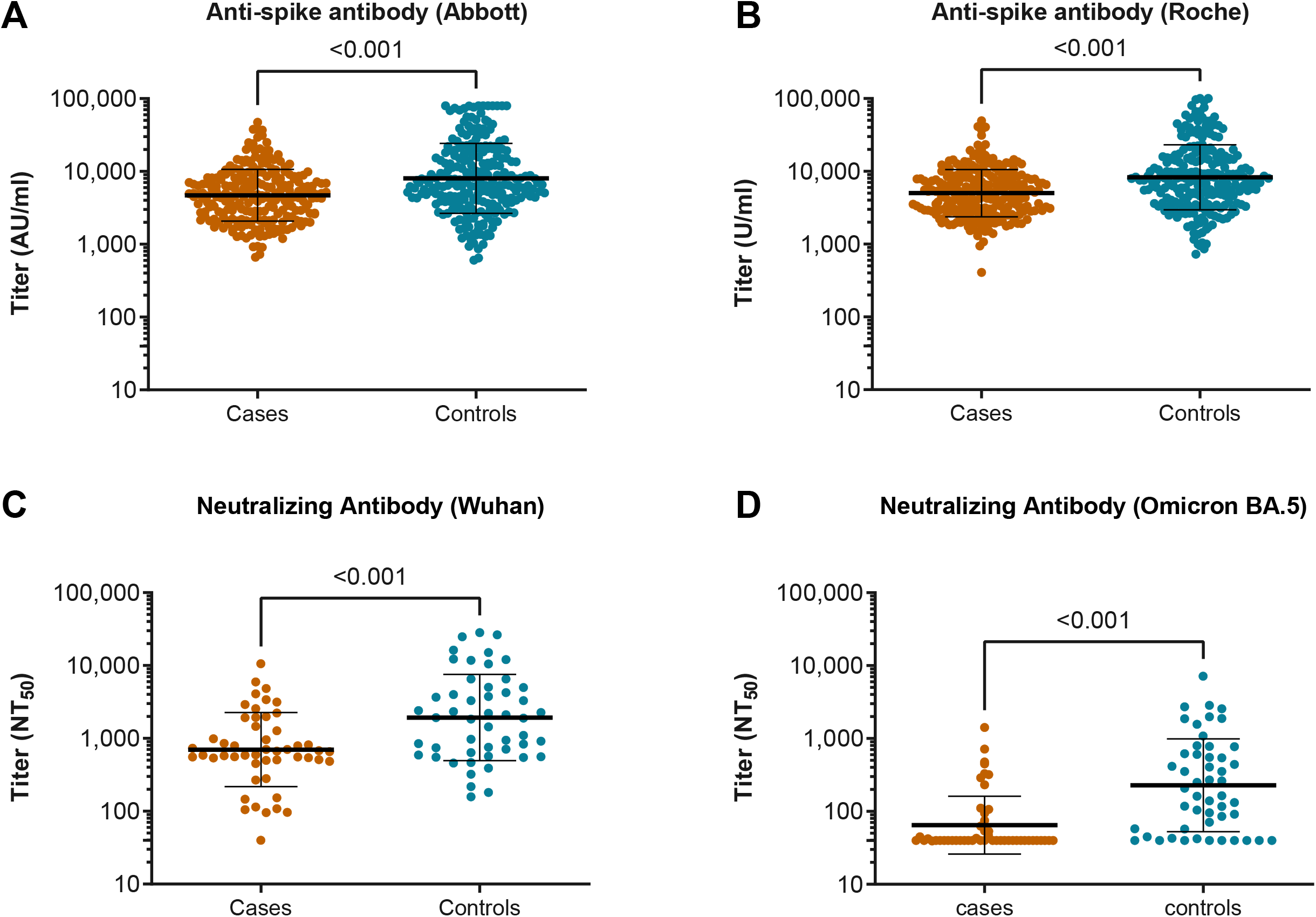
Comparison of the pre-infection neutralizing and anti-spike antibody titers between propensity-score matched cases and controls. Pre-infection anti-spike antibody titers measured using the Abbott reagent (**A**) and Roche reagent (**B**) among 243 cases with breakthrough infection and 243 matched controls. In addition, the neutralizing antibody titers against Wuhan (**C**) and Omicron BA.5 (**D**) among the 50 matched pairs were randomly selected from 243 matched pairs. In each panel, the horizontal bars indicate the geometric mean titers and the I-shaped bars indicate the geometric standard deviations. Abbreviations: AU, arbitrary units; NT_50_, 50% neutralizing titer

### Antibody Testing

Neutralizing activity against Wuhan and Omicron BA.5 in the sera of patients and controls was determined by quantifying the serum-mediated suppression of the cytopathic effect of each SARS-CoV-2 strain in HeLa_hACE2-TMPRSS2_ cells.^11, 12^ The details of the measurement methods are described in **eText 2**.

We assessed anti–SARS-CoV-2 antibodies in all the participants at baseline and retrieved data for the case-control pairs. We quantitatively measured the levels of antibodies against the receptor-binding domain of the SARS-CoV-2 spike protein using the AdviseDx SARS-CoV-2 IgG II assay (Abbott) (immunoglobulin [Ig] G) and Elecsys^®^ Anti-SARS-CoV-2 S RUO (Roche) (including IgG). We also qualitatively measured antibodies against the SARS-CoV-2 nucleocapsid (N) protein using the SARS-CoV-2 IgG assay (Abbott) and Elecsys^®^ Anti-SARS-CoV-2 RUO (Roche) to determine those with a possible infection before the baseline survey.

### Long COVID

We defined long COVID as reporting SARS-CoV-2-related symptoms for ≥4 weeks after the SARS-CoV-2 infection, according to the definition of the Centers for Disease Control and Prevention (CDC).^15^ We asked patients with SARS-CoV-2 infections the following question in a follow-up survey conducted in December 2022: “Have you had any symptoms persisting for 4 weeks (28 days) or more since you were infected with COVID-19?” The participants who answered yes were asked about their symptoms in detail. The participants were informed that symptoms persisting for 4 weeks while resolving or returning should also be included, whereas those that were apparently due to other illnesses should not be included. We created the following six categories for the analysis, in accordance with the CDC guidelines^15^: (1) any of the long COVID symptoms; (2) general symptoms (tiredness/fatigue or fever); (3) respiratory and cardiac symptoms (difficulty breathing, cough, chest pain, or heart palpitations); (4) neurological symptoms (difficulty thinking/concentrating, headache, sleep problems, changes in smell/taste, or depression/anxiety); (5) digestive symptoms (diarrhea or stomach pain); and (6) other symptoms (joint/muscle pain, rash, or others). Among the 243 cases with symptomatic breakthrough infections included in the case-control analysis, we included 166 cases who participated in the follow-up survey and answered the questionnaire regarding long COVID in the analysis of the association between pre-infection antibody titers and long COVID development.

### Statistical Analyses

We compared the log-transformed titers of neutralizing (Wuhan and Omicron BA.5) and anti-spike antibodies between matched pairs using a generalized estimating equation (GEE) with group assignment (case or control) and a robust variance estimator to examine the difference in pre-infection antibody levels between cases and controls. Next, we back-transformed and presented these values as geometric mean titers (GMTs) with 95% confidence intervals (CIs). We repeated the analysis by restricting those who were infection-naïve at baseline (no history of COVID-19 and negative anti-N assays at baseline) for sensitivity analysis. We compared the titers between infection-naïve, previously diagnosed infection (history of COVID-19), and previously undiagnosed infection (anti-SARS-CoV-2 N seropositive on Abbott or Roche assays at baseline without a history of COVID-19) using a linear regression model adjusted for age, sex, and the interval between vaccination and blood sampling to examine the association between previous SARS-CoV-2 infection status and baseline antibody titers. We ran a linear regression model with adjustments for age, sex, previous SARS-COV-2 infection status, and the interval between vaccination and blood sampling to examine the association between pre-infection antibody titers and long COVID. We used the Kruskal–Wallis test with Dunn’s multiple comparisons to compare the neutralizing ratio of Omicron BA.5 to Wuhan NT_50_ between those with and without previous infection. Statistical analyses were performed using Stata version 17.0 (StataCorp LLC), and graphics were generated using GraphPad Prism 9 (GraphPad, Inc.). All *P*-values were 2-sided, and the statistical significance was set at *P*<0.05.

## Results

### Baseline characteristics in the unmatched and matched cohorts

We ascertained 243 symptomatic breakthrough infection cases during the follow-up in the unmatched cohort of third-dose recipients, with an incidence rate of 13.0 per 10000 person-days. All patients had mild symptoms, and only three were admitted to the hospital. All 94 cases with available information on the SARS-CoV-2 strain type were Omicron variants. Of these, 2% were estimated to be Omicron BA.1; 5%, Omicron BA.2; and 85%, Omicron BA.4/BA.5, whereas the subvariants of the remaining 7% could not be determined. Patients were more likely to be younger, nurses, and at moderate or high risk of occupational SARS-CoV-2 exposure than were the controls in the unmatched cohort (**Table 1**). The 243 matched pairs were well-balanced regarding all the baseline characteristics after propensity matching with a 1:1 ratio.

**Table 1.**
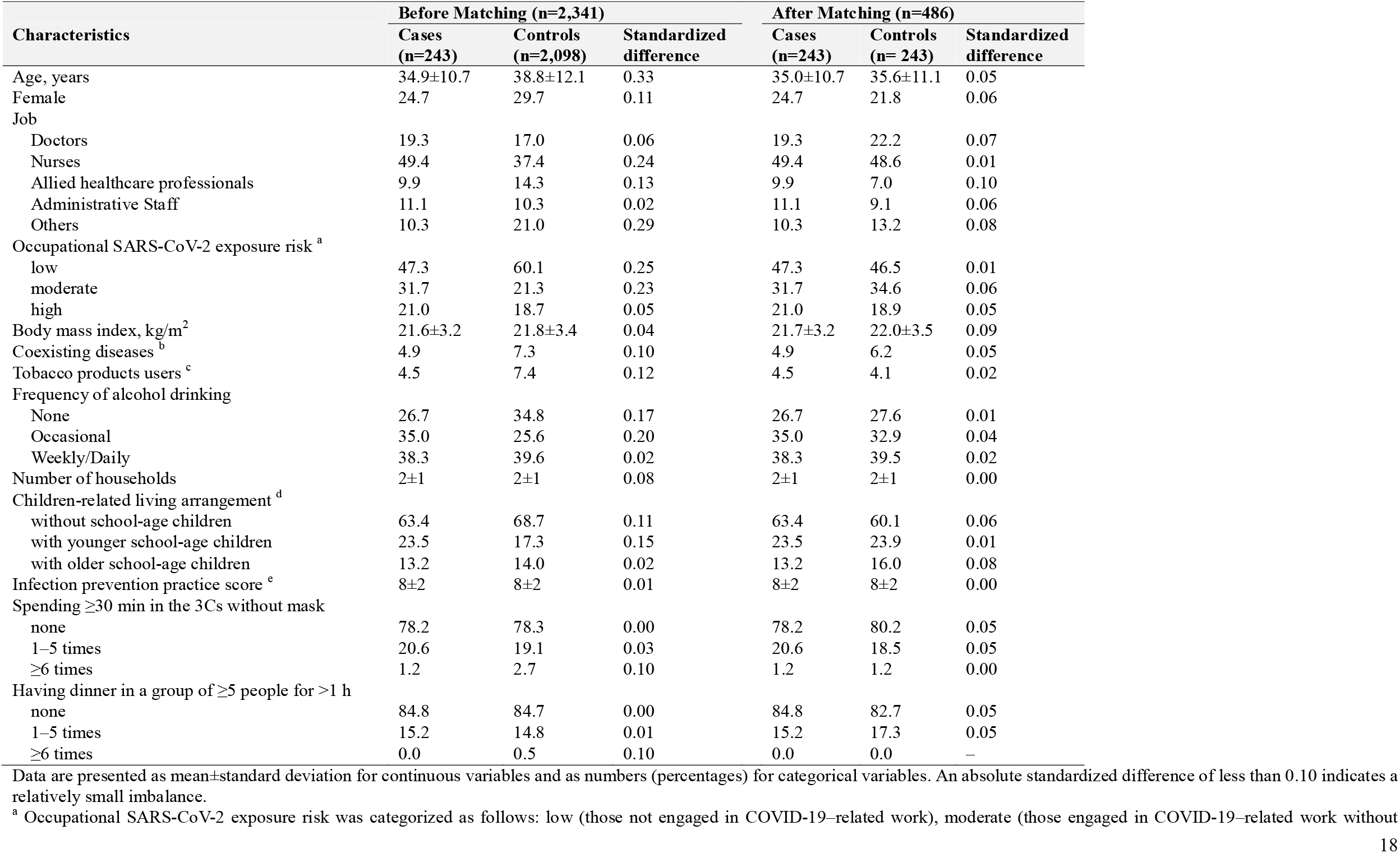
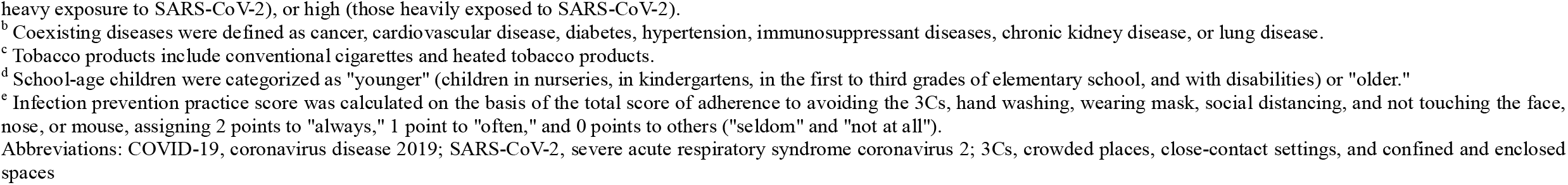
Baseline Characteristics Before and After Propensity Score Matching

| Characteristics | Before Matching (n=2,341) |  |  | After Matching (n=486) |  |  |
| --- | --- | --- | --- | --- | --- | --- |
|  | Cases (n=243) | Controls (n=2,098) | Standardized difference | Cases (n=243) | Controls (n= 243) | Standardized difference |
| Age, years | 34.9±10.7 | 38.8±12.1 | 0.33 | 35.0±10.7 | 35.6±11.1 | 0.05 |
| Female | 24.7 | 29.7 | 0.11 | 24.7 | 21.8 | 0.06 |
| Job |  |  |  |  |  |  |
| Doctors | 19.3 | 17.0 | 0.06 | 19.3 | 22.2 | 0.07 |
| Nurses | 49.4 | 37.4 | 0.24 | 49.4 | 48.6 | 0.01 |
| Allied healthcare professionals | 9.9 | 14.3 | 0.13 | 9.9 | 7.0 | 0.10 |
| Administrative Staff | 11.1 | 10.3 | 0.02 | 11.1 | 9.1 | 0.06 |
| Others | 10.3 | 21.0 | 0.29 | 10.3 | 13.2 | 0.08 |
| Occupational SARS-CoV-2 exposure risk <sup>a</sup> |  |  |  |  |  |  |
| low | 47.3 | 60.1 | 0.25 | 47.3 | 46.5 | 0.01 |
| moderate | 31.7 | 21.3 | 0.23 | 31.7 | 34.6 | 0.06 |
| high | 21.0 | 18.7 | 0.05 | 21.0 | 18.9 | 0.05 |
| Body mass index, kg/m<sup>2</sup> | 21.6±3.2 | 21.8±3.4 | 0.04 | 21.7±3.2 | 22.0±3.5 | 0.09 |
| Coexisting diseases <sup>b</sup> | 4.9 | 7.3 | 0.10 | 4.9 | 6.2 | 0.05 |
| Tobacco products users <sup>c</sup> | 4.5 | 7.4 | 0.12 | 4.5 | 4.1 | 0.02 |
| Frequency of alcohol drinking |  |  |  |  |  |  |
| None | 26.7 | 34.8 | 0.17 | 26.7 | 27.6 | 0.01 |
| Occasional | 35.0 | 25.6 | 0.20 | 35.0 | 32.9 | 0.04 |
| Weekly/Daily | 38.3 | 39.6 | 0.02 | 38.3 | 39.5 | 0.02 |
| Number of households | 2±1 | 2±1 | 0.08 | 2±1 | 2±1 | 0.00 |
| Children-related living arrangement <sup>d</sup> |  |  |  |  |  |  |
| without school-age children | 63.4 | 68.7 | 0.11 | 63.4 | 60.1 | 0.06 |
| with younger school-age children | 23.5 | 17.3 | 0.15 | 23.5 | 23.9 | 0.01 |
| with older school-age children | 13.2 | 14.0 | 0.02 | 13.2 | 16.0 | 0.08 |
| Infection prevention practice score <sup>e</sup> | 8±2 | 8±2 | 0.01 | 8±2 | 8±2 | 0.00 |
| Spending ≥30 min in the 3Cs without mask |  |  |  |  |  |  |
| none | 78.2 | 78.3 | 0.00 | 78.2 | 80.2 | 0.05 |
| 1–5 times | 20.6 | 19.1 | 0.03 | 20.6 | 18.5 | 0.05 |
| ≥6 times | 1.2 | 2.7 | 0.10 | 1.2 | 1.2 | 0.00 |
| Having dinner in a group of ≥5 people for >1 h |  |  |  |  |  |  |
| none | 84.8 | 84.7 | 0.00 | 84.8 | 82.7 | 0.05 |
| 1–5 times | 15.2 | 14.8 | 0.01 | 15.2 | 17.3 | 0.05 |
| ≥6 times | 0.0 | 0.5 | 0.10 | 0.0 | 0.0 | – |

Data are presented as mean±standard deviation for continuous variables and as numbers (percentages) for categorical variables. An absolute standardized difference of less than 0.10 indicates a relatively small imbalance.
<sup>a</sup> Occupational SARS-CoV-2 exposure risk was categorized as follows: low (those not engaged in COVID-19–related work), moderate (those engaged in COVID-19–related work without
heavy exposure to SARS-CoV-2), or high (those heavily exposed to SARS-CoV-2).
<sup>b</sup> Coexisting diseases were defined as cancer, cardiovascular disease, diabetes, hypertension, immunosuppressant diseases, chronic kidney disease, or lung disease.
<sup>c</sup> Tobacco products include conventional cigarettes and heated tobacco products.
<sup>d</sup> School-age children were categorized as "younger" (children in nurseries, in kindergartens, in the first to third grades of elementary school, and with disabilities) or "older."
<sup>e</sup> Infection prevention practice score was calculated on the basis of the total score of adherence to avoiding the 3Cs, hand washing, wearing mask, social distancing, and not touching the face, nose, or mouth, assigning 2 points to "always," 1 point to "often," and 0 points to others ("seldom" and "not at all").
Abbreviations: COVID-19, coronavirus disease 2019; SARS-CoV-2, severe acute respiratory syndrome coronavirus 2; 3Cs, crowded places, close-contact settings, and confined and enclosed spaces

Patients were less likely than controls to have had a previously diagnosed infection during the Omicron BA.1/BA.2 waves (1.2% vs. 11.1%) and a previously undiagnosed infection (2.9% vs. 8.2%) (**Table 2**). The interval between the third dose and blood sampling did not show a significant difference among the groups; the median intervals for cases and controls were 174 days (interquartile range [IQR]:153–184) and 173 days (IQR:153–183), respectively. The type of mRNA vaccine received did not differ between cases and controls, and most cases (91.4%) and controls (93.0%) received three doses of BNT162b2.

**Table 2.**
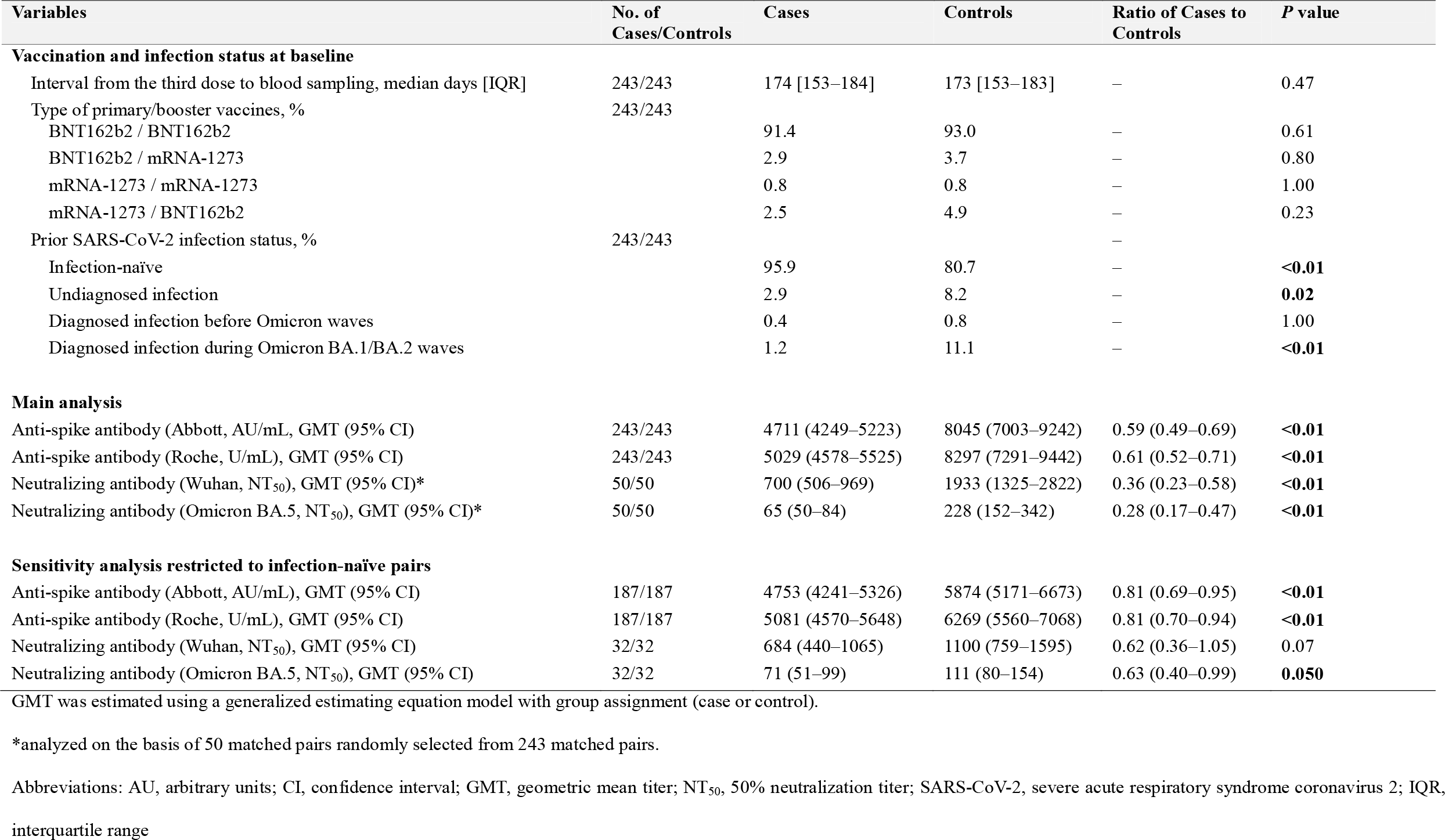
Comparison of Pre-infection Antibody Titers, Intervals Since the Third Vaccine Dose, and Prior Infection Status Between Cases and Controls

| Variables | No. of Cases/Controls | Cases | Controls | Ratio of Cases to Controls | P value |
| --- | --- | --- | --- | --- | --- |
| **Vaccination and infection status at baseline** |  |  |  |  |  |
| Interval from the third dose to blood sampling, median days [IQR] | 243/243 | 174 [153–184] | 173 [153–183] | – | 0.47 |
| Type of primary/booster vaccines, % | 243/243 |  |  |  |  |
| BNT162b2 / BNT162b2 |  | 91.4 | 93.0 | – | 0.61 |
| BNT162b2 / mRNA-1273 |  | 2.9 | 3.7 | – | 0.80 |
| mRNA-1273 / mRNA-1273 |  | 0.8 | 0.8 | – | 1.00 |
| mRNA-1273 / BNT162b2 |  | 2.5 | 4.9 | – | 0.23 |
| Prior SARS-CoV-2 infection status, % | 243/243 |  |  | – |  |
| Infection-naïve |  | 95.9 | 80.7 | – | **<0.01** |
| Undiagnosed infection |  | 2.9 | 8.2 | – | **0.02** |
| Diagnosed infection before Omicron waves |  | 0.4 | 0.8 | – | 1.00 |
| Diagnosed infection during Omicron BA.1/BA.2 waves |  | 1.2 | 11.1 | – | **<0.01** |
| **Main analysis** |  |  |  |  |  |
| Anti-spike antibody (Abbott, AU/mL, GMT (95% CI) | 243/243 | 4711 (4249–5223) | 8045 (7003–9242) | 0.59 (0.49–0.69) | **<0.01** |
| Anti-spike antibody (Roche, U/mL), GMT (95% CI) | 243/243 | 5029 (4578–5525) | 8297 (7291–9442) | 0.61 (0.52–0.71) | **<0.01** |
| Neutralizing antibody (Wuhan, NT<sub>50</sub>), GMT (95% CI)* | 50/50 | 700 (506–969) | 1933 (1325–2822) | 0.36 (0.23–0.58) | **<0.01** |
| Neutralizing antibody (Omicron BA.5, NT<sub>50</sub>), GMT (95% CI)* | 50/50 | 65 (50–84) | 228 (152–342) | 0.28 (0.17–0.47) | **<0.01** |
| **Sensitivity analysis restricted to infection-naïve pairs** |  |  |  |  |  |
| Anti-spike antibody (Abbott, AU/mL), GMT (95% CI) | 187/187 | 4753 (4241–5326) | 5874 (5171–6673) | 0.81 (0.69–0.95) | **<0.01** |
| Anti-spike antibody (Roche, U/mL), GMT (95% CI) | 187/187 | 5081 (4570–5648) | 6269 (5560–7068) | 0.81 (0.70–0.94) | **<0.01** |
| Neutralizing antibody (Wuhan, NT<sub>50</sub>), GMT (95% CI) | 32/32 | 684 (440–1065) | 1100 (759–1595) | 0.62 (0.36–1.05) | 0.07 |
| Neutralizing antibody (Omicron BA.5, NT<sub>50</sub>), GMT (95% CI) | 32/32 | 71 (51–99) | 111 (80–154) | 0.63 (0.40–0.99) | **0.050** |

GMT was estimated using a generalized estimating equation model with group assignment (case or control).
\*analyzed on the basis of 50 matched pairs randomly selected from 243 matched pairs.
Abbreviations: AU, arbitrary units; CI, confidence interval; GMT, geometric mean titer; NT<sub>50</sub>, 50% neutralization titer; SARS-CoV-2, severe acute respiratory syndrome coronavirus 2; IQR, interquartile range

### Correlation between anti-spike and neutralizing antibodies

Anti-spike antibody titers with the Abbott and Roche assays highly correlated with neutralizing antibody titers against Wuhan (Spearman’s ρ:0.82 and 0.84, respectively) and those against Omicron BA.5 (Spearman’s ρ:0.80 and 0.85, respectively) (**eFigure 1**).

### Pre-infection antibody titers between the matched cases and controls

Pre-infection anti-spike and neutralizing antibody titers were lower in patients than in controls. The GEE-predicted GMTs (95% CI) of the anti-spike antibody on Abbott assay (AU/ml) was 4711 (4249–5223) for cases and 8045 (7003–9242) for controls with a predicted case-to-control ratio of the titers of 0.59 (95% CI:0.49–0.69) (**Table 2** and **Figure 1**). The GMTs (95% CI) of the anti-spike antibody on Roche assay (U/mL) were 5029 (4578–5525) for cases and 8297 (7291–9442) with a ratio of 0.61 (95% CI:0.52–0.71). The predicted neutralizing antibody GMTs (95% CI) against Wuhan (NT_50_) were 700 (506–969) for cases and 1933 (1325–2822) for controls, with a ratio of 0.36 (95% CI:0.23–0.58). Those against Omicron BA.5 were 65 (50–84) for cases and 228 (152–342) for controls, with a ratio of 0.28 (95% CI:0.17–0.47). The difference in pre-infection antibody titers between cases and controls was attenuated in the sensitivity analysis restricting cases and controls to infection-naïve pairs; however, titers were still statistically lower in cases, except for neutralizing titers against Wuhan (**Table 2**).

### Previous SARS-CoV-2 infection status and antibodies

Baseline antibody titers were higher in previously diagnosed and undiagnosed infected individuals than in infection-naïve individuals, whereas the diagnosed and undiagnosed groups did not show significant differences in the titers (**Figures 2A-D**). Compared with infection-naïve individuals, diagnosed and undiagnosed infected individuals had 4.8- and 4.4-fold higher anti-spike antibody titers using the Abbott assay, 4.2- and 3.5-fold higher titers using the Roche assay, 8.2- and 4.5-fold higher neutralizing titers against Wuhan, and 12.8- and 6.7-fold higher titers against Omicron BA.5, respectively.

**Figure 2.**
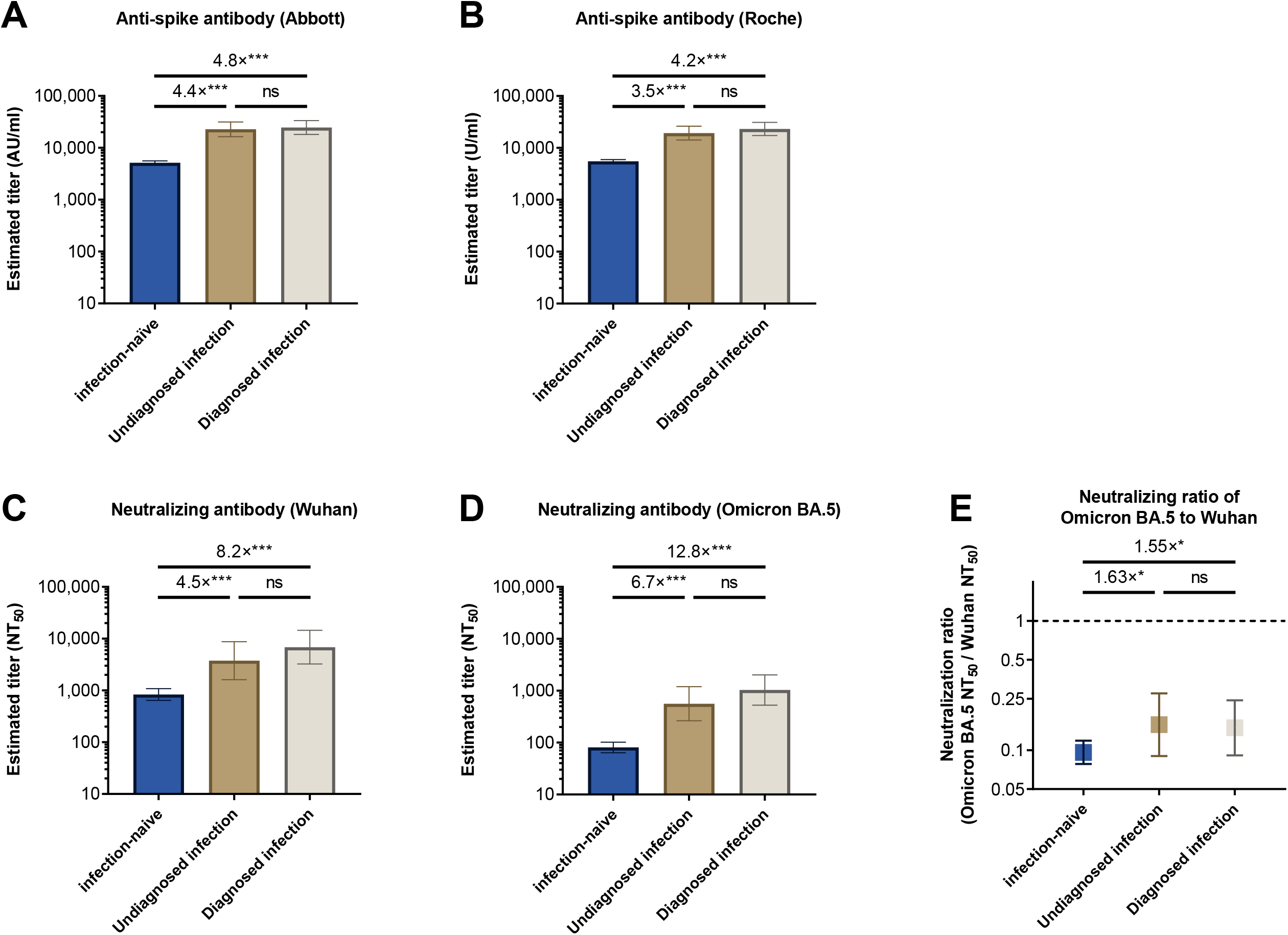
Association between previous SARS-CoV-2 infection status and anti-spike and neutralizing antibodies at baseline A diagnosed infection was defined as a history of COVID-19, whereas an undiagnosed infection was defined as anti-SARS-CoV-2 nucleocapsid seropositivity using Abbott or Roche reagents at baseline. (**A-D**) Associations of previous SARS-CoV-2 infection status with anti-spike antibody titers using the Abbott (A) and Roche (B) reagents and neutralizing antibody titers against Wuhan (C) and Omicron BA.5 (D). The bars indicate geometric mean antibody titers and I-shaped bars indicate their confidence intervals, estimated using a multivariate linear regression model with adjustment for age, sex, and the interval between the third vaccination and blood sampling. (**E**) The neutralizing ratio of Omicron BA.5 to Wuhan across the previous SARS-CoV-2 infection status. Error bars indicate geometric mean ratios with 95% confidence intervals. The dotted line indicates that the neutralization of Wuhan and Omicron BA.5 was equal. The nonparametric Kruskal–Wallis test with Dunn’s multiple comparison correction was used to compare the ratios across groups. Abbreviations: COVID-19, coronavirus disease 2019; SARS-CoV-2, severe acute respiratory syndrome coronavirus 2; AU, arbitrary units; NT_50_, 50% neutralizing titer; ns, not significant; * *P*<0.05; ** *P*<0.01; *** *P*<0.001.

The neutralizing antibodies titers against Omicron BA.5 were considerably lower than those against Wuhan in the total samples, with a GMT ratio of 0.10 for Omicron BA.5 to Wuhan (95% CI:0.08–0.13). The GMT ratio was greater for diagnosed (0.15, 95% CI:0.09–0.24) and undiagnosed (0.16, 95% CI:0.09–0.28) infection groups than for the infection-naïve group (0.10, 95% CI:0.08–0.12) (**Figure 2E**).

### Pre-infection antibody titers and long COVID

The proportion of long COVID cases was 26.5% (95% CI:20.0–33.9). Respiratory and cardiac (18.7%), general (12.7%), and neurological (7.8%) symptoms were the most frequent. None of the patients reported digestive symptoms.

Pre-infection antibody titers did not show material differences between those who reported any of the long-term COVID symptoms and those who did not (**Table 3**). Regarding the long-COVID symptoms, general, respiratory, cardiac, neurological, gastrointestinal, and other symptoms did not show differences between the groups.

**Table 3.** Association of Pre-infection Anti-spike and Neutralizing Antibody Titers With Long-COVID Developments Among Breakthrough Infection Cases

| Long-COVID | N | Anti-spike antibody<br>(Abbott), AU/ml | Anti-spike antibody<br>(Roche), U/ml | N | NAb against<br>Wuhan, NT<sub>50</sub> | NAb against<br>Omicron BA.5, NT<sub>50</sub> |
| --- | --- | --- | --- | --- | --- | --- |
|  |  | Ratio of mean (95% CI) | Ratio of mean (95% CI) |  | Ratio of mean (95% CI) | Ratio of mean (95% CI) |
| Any of long-COVID symptoms |  |  |  |  |  |  |
| No | 122 | Reference | Reference | 22 | Reference | Reference |
| Yes | 44 | 1.23 (0.95–1.60) | 1.17 (0.92–1.48) | 11 | 1.53 (0.56–4.23) | 1.04 (0.50–2.14) |
| General symptoms <sup>a</sup> |  |  |  |  |  |  |
| No | 145 | Reference | Reference | 30 | Reference | Reference |
| Yes | 21 | 1.15 (0.82–1.62) | 1.11 (0.81–1.52) | 3 | 2.13 (0.36–12.4) | 1.04 (0.29–3.70) |
| Respiratory and heart symptoms <sup>b</sup> |  |  |  |  |  |  |
| No | 135 | Reference | Reference | 24 | Reference | Reference |
| Yes | 31 | 1.14 (0.85–1.53) | 1.13 (0.87–1.48) | 9 | 1.52 (0.54–4.28) | 1.03 (0.49–2.15) |
| Neurological symptoms <sup>c</sup> |  |  |  |  |  |  |
| No | 153 | Reference | Reference | 32 | Reference | Reference |
| Yes | 13 | 1.21 (0.79–1.84) | 1.19 (0.81–1.75) | 1 | 0.84 (0.05–14.4) | 0.55 (0.07–4.00) |
| Digestive symptoms <sup>d</sup> |  |  |  |  |  |  |
| No | 166 | Reference | Reference | 33 | Reference | Reference |
| Yes | 0 | NA | NA | 0 | NA | NA |
| Other symptoms <sup>e</sup> |  |  |  |  |  |  |
| No | 157 | Reference | Reference | 30 | Reference | Reference |
| Yes | 9 | 1.56 (0.94–2.59) | 1.23 (0.77–1.96) | 3 | 2.39 (0.42–13.5) | 1.75 (0.51–5.98) |

Data are shown as the ratio of means estimated using the multivariate linear regression model adjusted for age, sex, interval between the third vaccination and blood sampling, and previous SARS-CoV-2 infection status.
<sup>a</sup> tiredness/fatigue or fever.
<sup>b</sup> difficulty breathing, cough, chest pain, or heart palpitations
<sup>c</sup> difficulty thinking/concentrating, headache, sleep problems, changes in smell/taste, or depression/anxiety
<sup>d</sup> diarrhea or stomach pain
<sup>e</sup> joint/muscle pain, rash, or others
Abbreviations: AU, arbitrary units; CI, confidence interval; COVID, coronavirus disease; SARS-CoV-2, severe acute respiratory syndrome coronavirus 2; NA, not applicable

## Discussion

The pre-infection neutralizing antibodies against Omicron BA.5 were lower in infected cases during the Omicron BA.5 wave than in the matched controls in this nested case-control study of a cohort of healthcare workers approximately 6 months after the third vaccination. The pre-infection neutralizing capacity did not show material differences between the breakthrough cases who reported long COVID and those who did not. This is the first study to investigate the association of pre-infection neutralizing antibody titers with the risk of Omicron BA.5 infection and development of long COVID.

The significant association of pre-infection antibody titers with Omicron BA.5 infection in this study contrasts with the findings of our previous studies, where no associations were found with the risk of Delta infection among individuals approximately 2 months after the second dose^3^ and with the risk of Omicron BA.1/BA.2 infection among those approximately 1 month after the third dose.^4^ This may be attributed to the difference in variability of humoral immunity levels at the time of blood sampling across the surveys. Specifically, the coefficient of variation (CV) of anti-spike (Abbott) and Wuhan neutralizing antibody titers in blood samples in the present study (133% and 233%, respectively) were larger than those for Delta (69% and 69%, respectively) and Omicron BA.1/BA.2 (56% and 90%, respectively) infections (**eTable 3**). Among the third-dose recipients without a history of COVID-19, those approximately 6 months after vaccination (present study) had 4.5-fold lower mean antibody titers (Abbott) but a 1.9-fold greater CV than those approximately 1 month after vaccination (data from a previous study on Omicron BA.1/BA.2 infection). These results are compatible with longitudinal studies showing a large variation in the speed of waning of anti-SARS-CoV-2 antibodies across vaccine recipients with different characteristics.^16-18^ Antibody levels can be used as a predictor for infection risk in populations with large variations in humoral immunity across individuals.

In this study, those with previous SARS-CoV-2 infection (mostly Omicron BA.1/BA.2) among three-dose vaccine recipients had higher neutralizing antibody titers against Omicron BA.5 and a greater neutralizing ratio of Omicron BA.5 to Wuhan than those who were infection-naïve, in agreement with the findings of previous studies.^18, 19^ The proportion of previous Omicron BA.1/BA.2 infection was higher in controls than in breakthrough cases, consistent with the results of a previous study.^20^ These results suggest that the Omicron BA.5 neutralizing capacity induced by previous Omicron BA.1/BA.2 infections lowers the risk of Omicron BA.5 infection among the three-dose recipients and is compatible with the findings of observational studies indicating that hybrid immunity (three vaccinations plus previous Omicron BA.1/BA.2 infection) has higher effectiveness against Omicron BA.5 infection than only three vaccinations.^20^ We confirmed that Omicron BA.5 neutralizing antibody titers were modestly but significantly lower in cases than in controls in the analysis of infection-naïve vaccine recipients, suggesting that the level of humoral immunity induced by historical mRNA vaccine alone can also predict, albeit to a lesser extent than hybrid immunity, the infection risk of Omicron BA.5 with a high immune-evasive nature.

In this study, 26.5% (95% CI:20.0–33.9) of patients who received three vaccinations and were infected during the Omicron BA.5-predominate wave experienced long COVID, which is somewhat higher the proportion among US adults who were infected with Omicron BA.4/BA.5 after receiving the third vaccine dose (20.9%, 95% CI:16.4–26.2).^21^ We found no association between pre-infection antibody titers and the risk of long COVID. An Itarian study reported that anti-spike IgG titers measured during the acute infection phase did not predict long COVID in vaccinated patients with or without hospitalization.^22^ Evidence for the association between vaccination status and long COVID risk is also inconsistent.^21, 23^ Our results and previous reports^21-23^ suggest that vaccine-induced immunity has no apparent protective role against post–COVID-19 symptoms.

This study had several strengths. We rigorously matched cases and controls using a propensity score estimated by several factors potentially associated with SARS-CoV-2 infection risk, including occupational SARS-CoV-2 exposure risk, living arrangements, comorbidities, infection prevention practices, and high infection risk–behaviors. Blood samples for antibody testing were obtained before infection (1 month before the Omicron BA.5 epidemic onset). Previous SARS-CoV-2 infection was determined according to the history of COVID-19 infection and results of anti-N assay tests, allowing us to identify undiagnosed infections. We measured the neutralizing antibody titers against Wuhan and Omicron BA.5 using live viruses.

This study had some limitations. We did not conduct active surveillance to detect SARS-CoV-2 infection during the follow-up period. Nonetheless, we confirmed that the results were virtually unchanged in the sensitivity analysis, which excluded individuals who tested seropositive in anti-N assays in the follow-up survey from the control group (**eTable 2**). Data on virus strain were available for 39% of the cases; however, the remaining cases of breakthrough infections were most likely due to the Omicron BA.5 variant, which accounted for more than 90% of sequenced COVID-19 samples in Japan during the follow-up (July to September 2022).^24^ Lower levels of pre-infection humoral immunity have been linked to severe forms of COVID-19,^25, 26^ which may increase the risk of long COVID.^27^ Our results regarding long-COVID symptoms cannot be applied to patients with severe symptoms, since all the included patients with COVID-19 had mild symptoms. The sample size for the analysis of the relationship between antibody titers and long COVID (n=166) may be insufficient to detect a significant effect.

## Conclusion

Pre-infection and live-virus neutralizing antibody titers against Omicron BA.5 were lower in breakthrough infection cases than in their matched controls during the Omicron BA.5– dominant wave among the third-dose recipients (mainly three doses of the BNT162b vaccine) approximately 6 months post-vaccination. The high neutralizing capacity of individuals with a history of Omicron BA.1/BA.2 infection was a substantial cause of these differences. Pre-infection neutralizing antibody titers were not associated with the risk of long COVID.

## Data Availability

All data produced in the present study are available upon reasonable request to the authors.

## Author Contributions

Drs. Yamamoto and Mizoue had full access to all data in the study and took responsibility for the integrity of the data and the accuracy of the data analysis.

Drs Yamamoto and Matsuda contributed equally to this article.

*Concept and design*: Yamamoto S, Matsuda K, Maeda K, Mizoue T, Sugiyama H, Mitsuya H, Sugiura W, Ohmagari N.

*Acquisition, analysis, or interpretation of data*: Yamamoto S, Matsuda K, Maeda K, Horii K, Okudera K, Oshiro Y, Inamura N, Nemoto T, Takeuchi S. J, Li Y, Konishi M, Ozeki M, Mizoue T.

*Drafting of the manuscript*: Yamamoto S, Matsuda K, Mizoue T.

*Critical revision of the manuscript for important intellectual content*: Yamamoto S Matsuda K, Maeda K, Takeuchi S. J, Mizoue T, Sugiyama H, Aoyanagi N, Mitsuya H, Sugiura W, Ohmagari N.

*Statistical analysis*: Yamamoto S, Matsuda K, Konishi M.

*Administrative, technical, or material support*: Yamamoto S, Matsuda K, Maeda K, Horii K, Okudera K, Oshiro Y, Inamura N, Nemoto T, Takeuchi S. J, Konishi M, Ozeki M, Tsuchiya K, Gatanaga H, Oka S, Mizoue T, Sugiyama H, Aoyanagi N, Mitsuya H, Sugiura W, Ohmagari N.

*Supervision*: Mizoue T, Ohmagari N.

## Additional Contributions

We thank Mika Shichishima for her contribution to data collection and the staff of the Laboratory Testing Department for their contribution to antibody testing.

