## Supplementary materials for "Pre-infection neutralizing antibodies, Omicron BA.5 breakthrough infection, and long COVID: a propensity score-matched analysis"

**Supplemental Materials**

**eText 1. Propensity score matching algorithm**

We performed propensity score matching using a 1:1 matching algorithm without replacement, with a caliper width equal to 0.2 of the standard deviation of the logit of the propensity score, to identify controls with baseline characteristics similar to those of the patients. We estimated the propensity score using multivariable logistic regression, with the incidence of symptomatic SARS-CoV-2 infection as the dependent variable and the following variables as covariates: sex, age, job, occupational risk of SARS-CoV-2 infection, body mass index, coexisting diseases (cancer, cardiovascular diseases, diabetes, hypertension, immunosuppressive diseases, chronic kidney disease, and lung diseases), several infection prevention/risk behaviors, use of tobacco products, frequency of alcohol consumption, number of household members, and children-related living arrangements. Absolute standardized differences were estimated for all baseline covariates before and after matching to assess the pre-match and post-match balances. Standardized differences <0.1 for a given covariate indicated a relatively small imbalance.

**eText 2.** **Measurement methods for neutralizing antibodies**

The neutralizing activity in the sera of cases and controls was determined by quantifying the serum-mediated suppression of the cytopathic effect (CPE) of each SARS-CoV-2 strain in HeLa_hACE2-TMPRSS2_ cells obtained from the Japanese Collection of Research Bioresources (JCRB) Cell Bank (Osaka, Japan).^11, 12^ Each serum sample was serially diluted five-fold in a culture medium. The diluted sera were incubated with 100 50% tissue culture infectious dose (TCID_50_) of the virus at 37°C for 20 min (final serum dilution range of 1:40 to 1:125,000). Next, the serum–virus mixtures were inoculated with HeLa_hACE2-TMPRSS2_ cells (1.0×10^4^/well) in 96-well plates. The SARS-CoV-2 strains used in these assays were as follows: a Wuhan wild-type strain (SARS-CoV-2^05-2N^)^13^ and an Omicron BA.5 variant (SARS-CoV-2^TKYS14631/2022^, GISAID Accession ID; EPI_ISL_12812500.1).^14^ The levels of CPE observed in SARS-CoV-2–exposed cells were determined using the WST-8 assay with the Cell Counting Kit-8 (Dojindo, Kumamoto, Japan) after culturing the cells for 3 days. The serum dilution that resulted in 50% inhibition of CPE was defined as the 50% neutralization titer (NT_50_). Each serum sample was tested in duplicate, and the average value was used for analysis. Laboratory technicians were blinded to the case-control status.

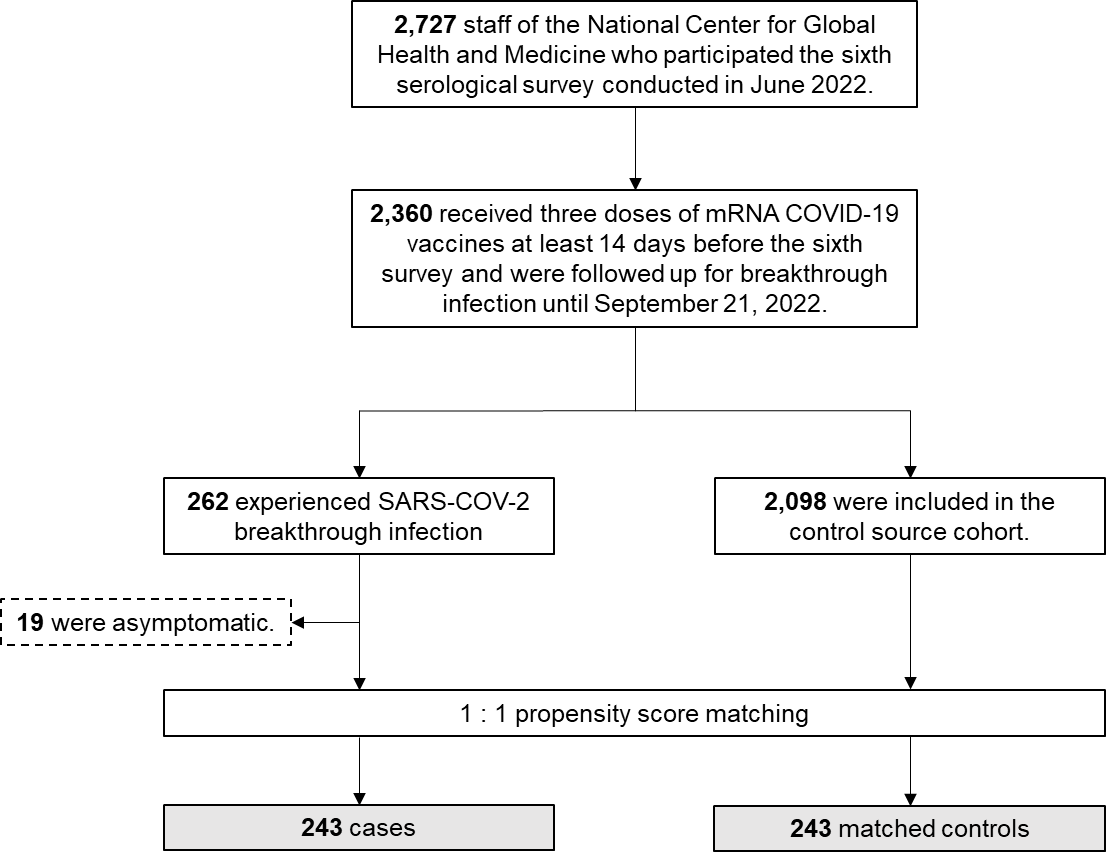

**eFigure 1. Case-control selection**

Abbreviations: COVID-19, coronavirus disease 2019; SARS-CoV-2, severe acute respiratory syndrome coronavirus 2

**eTable 1. Baseline characteristics of randomly selected and non-selected participants**

|  | **Total** |  |  | **Controls** |  |  | **Cases** |  |
| --- | --- | --- | --- | --- | --- | --- | --- | --- |
|  | **Non-selected** | **Selected** |  | **Non-selected** | **Selected** |  | **Non-selected** | **Selected** |
|  | N=386 | N=100 |  | N=193 | N=50 |  | N=193 | N=50 |
| Age, years | 32 [25–44] | 34.5 [26.5–45] |  | 32 [26–44] | 32 [25–45] |  | 31 [25–43] | 35.5 [27–45] |
| Female | 295 (76.4) | 78 (78.0) |  | 148 (76.7) | 42 (84.0) |  | 147 (76.2) | 36 (72.0) |
| Job |  |  |  |  |  |  |  |  |
| Doctors | 81 (21.0) | 20 (20.0) |  | 45 (23.3) | 9 (18.0) |  | 36 (18.7) | 11 (22.0) |
| Nurses | 192 (49.7) | 46 (46.0) |  | 93 (48.2) | 25 (50.0) |  | 99 (51.3) | 21 (42.0) |
| Allied health professionals | 35 ( 9.1) | 6 ( 6.0) |  | 15 ( 7.8) | 2 ( 4.0) |  | 20 (10.4) | 4 ( 8.0) |
| Addminisirative staff | 39 (10.1) | 10 (10.0) |  | 17 ( 8.8) | 5 (10.0) |  | 22 (11.4) | 5 (10.0) |
| Others | 39 (10.1) | 18 (18.0) |  | 23 (11.9) | 9 (18.0) |  | 16 ( 8.3) | 9 (18.0) |
| Occupational SARS-CoV-2 exposure risk | |  |  |  |  |  |  |  |
| Low | 176 (45.6) | 52 (52.0) |  | 88 (45.6) | 25 (50.0) |  | 88 (45.6) | 27 (54.0) |
| Moderate | 136 (35.2) | 25 (25.0) |  | 69 (35.8) | 15 (30.0) |  | 67 (34.7) | 10 (20.0) |
| High | 74 (19.2) | 23 (23.0) |  | 36 (18.7) | 10 (20.0) |  | 38 (19.7) | 13 (26.0) |
| Body mass index | 21.2 [19.5-23.2] | 21.0 [19.5-23.0] |  | 21.4 [19.7-23.6] | 20.9 [19.5-23.2] |  | 20.7 [19.4-22.9] | 21.0 [19.8-22.7] |
| Coexisting diseases | 22 ( 5.7) | 5 ( 5.0) |  | 12 ( 6.2) | 3 ( 6.0) |  | 10 ( 5.2) | 2 ( 4.0) |
| Tobacco products users | 18 ( 4.7) | 3 ( 3.0) |  | 9 ( 4.7) | 1 ( 2.0) |  | 9 ( 4.7) | 2 ( 4.0) |
| Frequency of alcohol drinking |  |  |  |  |  |  |  |  |
| Non | 104 (26.9) | 28 (28.0) |  | 53 (27.5) | 14 (28.0) |  | 51 (26.4) | 14 (28.0) |
| Occasional | 128 (33.2) | 37 (37.0) |  | 64 (33.2) | 16 (32.0) |  | 64 (33.2) | 21 (42.0) |
| Weekly/Daily | 154 (39.9) | 35 (35.0) |  | 76 (39.4) | 20 (40.0) |  | 78 (40.4) | 15 (30.0) |
| Number of households | 2 (1-3) | 3 (1-4) |  | 2 (1-4) | 2 (1-4) |  | 2 (1-3) | 3 (2-4) |
| Children-related living arrangement |  |  |  |  |  |  |  |  |
| Without school-age children | 250 (64.8) | 50 (50.0) |  | 117 (60.6) | 29 (58.0) |  | 133 (68.9) | 21 (42.0) |
| With younger school-age children | 81 (21.0) | 34 (34.0) |  | 44 (22.8) | 14 (28.0) |  | 37 (19.2) | 20 (40.0) |
| With older school-age children | 55 (14.2) | 16 (16.0) |  | 32 (16.6) | 7 (14.0) |  | 23 (11.9) | 9 (18.0) |
| Infection prevention practice score | 8 (7–10) | 8 (7–9) |  | 8 (7–10) | 8 (7–9) |  | 8 (7–10) | 8 (7–9) |
| Spending ≥30 min in the 3Cs without mask | |  |  |  |  |  |  |  |
| None | 304 (78.8) | 81 (81.0) |  | 153 (79.3) | 42 (84.0) |  | 151 (78.2) | 39 (78.0) |
| 1–5 times | 77 (19.9) | 18 (18.0) |  | 38 (19.7) | 7 (14.0) |  | 39 (20.2) | 11 (22.0) |
| ≥6 times | 5 ( 1.3) | 1 ( 1.0) |  | 2 ( 1.0) | 1 ( 2.0) |  | 3 ( 1.6) | 0 ( 0.0) |
| Having dinner in a group of ≥5 people for >1 h | |  |  |  |  |  |  |  |
| None | 321 (83.2) | 86 (86.0) |  | 155 (80.3) | 46 (92.0) |  | 166 (86.0) | 40 (80.0) |
| 1–5 times | 65 (16.8) | 14 (14.0) |  | 38 (19.7) | 4 ( 8.0) |  | 27 (14.0) | 10 (20.0) |
| ≥6 times | 0 | 0 |  | 0 | 0 |  | 0 | 0 |

Data are presented as median [interquartile range] or number (percentage).

Abbreviations: SARS-CoV-2, severe acute respiratory syndrome coronavirus 2

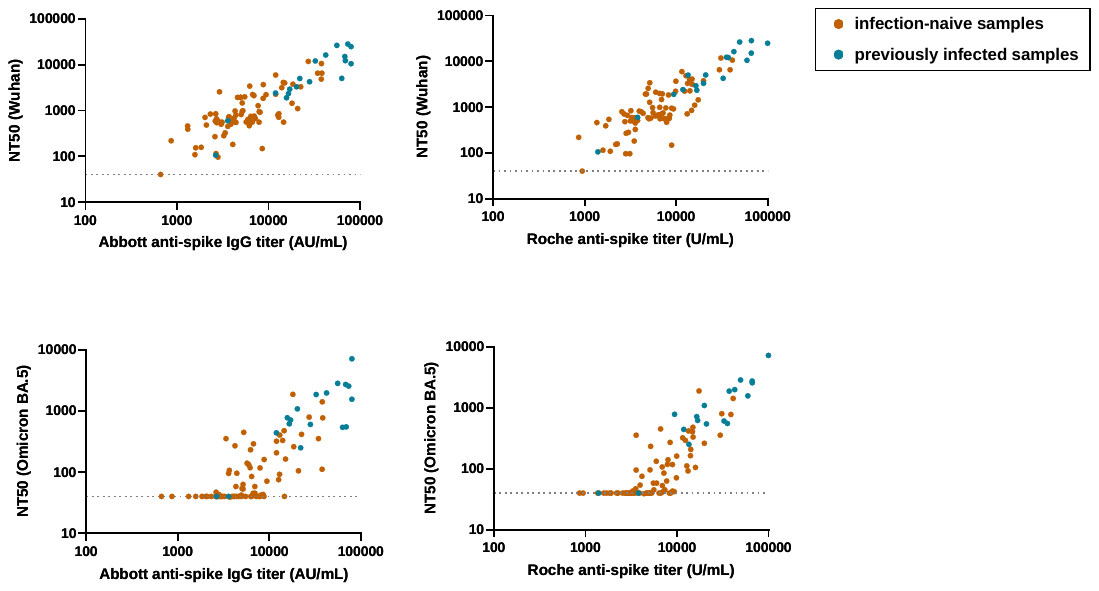

| **Spearman’s correlations** | **Total samples**  **(N=100)** | **Infection-naive samples**  **(N=82)** | **Previously infected samples**  **(N=18)** |
| --- | --- | --- | --- |
|  | ρ (95% CI) | ρ (95% CI) | ρ (95% CI) |
| **Abbott anti-spike IgG titer** |  |  |  |
| NT against Wuhan | 0.82 (0.74–0.87)*** | 0.71 (0.58–0.81)*** | 0.88 (0.70–0.97)*** |
| NT against Omicron BA.5 | 0.80 (0.71–0.86)*** | 0.67 (0.53–0.78)*** | 0.67 (0.27–0.87)** |
| **Roche anti-spike titer** |  |  |  |
| NT against Wuhan | 0.84 (0.76–0.89)*** | 0.74 (0.62–0.83)*** | 0.92 (0.80–0.97)*** |
| NT against Omicron BA.5 | 0.85 (0.78–0.90)*** | 0.75 (0.64–0.84)*** | 0.85 (0.62–0.94)*** |

****P*<0.001, ***P*<0.01, **P*<0.05

**eFigure 1. Correlation between anti-spike antibody titers and neutralizing antibody titers across previous SARS-CoV-2 infection status**

**eTable 2. Sensitivity analysis for the comparison of pre-infection antibody titers between cases and controls**

| **Variables** | **No. of Cases/Controls** | **Cases** | **Controls** | **Ratio of Cases to Controls** | ***P* value** |
| --- | --- | --- | --- | --- | --- |
| **Sensitivity analysis excluding those who became anti-N seropositive at follow-up survey from controls** |  |  |  |  |  |
| Anti-spike antibody (Abbott, AU/mL, GMT (95% CI) | 243/208 | 4711 (4249–5223) | 7046 (6114–8121) | 0.67 (0.56–0.79) | **<0.01** |
| Anti-spike antibody (Roche, U/mL), GMT (95% CI) | 243/208 | 5029 (4578–5525) | 7347 (6437–8385) | 0.68 (0.58–0.80) | **<0.01** |
| Neutralizing antibody (Wuhan, NT_50_), GMT (95% CI) | 50/40 | 700 (506–969) | 1445 (981–2130) | 0.48 (0.30–0.78) | **<0.01** |
| Neutralizing antibody (Omicron BA.5, NT_50_), GMT (95% CI) | 50/40 | 65 (50–84) | 161 (106–244) | 0.40 (0.24–0.68) | **<0.01** |

GMT was estimated using a generalized estimating equation model.

Abbreviations: AU, arbitrary units; CI, confidence interval; GMT, geometric mean titer; NT_50_, 50% neutralization titer; SARS-CoV-2, severe acute respiratory syndrome coronavirus 2

**eTable 3. Geometric coefficient of variation of the anti-spike and neutralizing antibody titers measured in present and previous case-control studies**

| **Case-control studies** | **Vaccination status** | **Previous infection status** | **Interval from last vaccination to blood sampling,**  **median days (IQR)** | **Anti-spike antibody (Abbott)** | | |  | **Anti-spike antibody (Roche)** | | |  | **Neutralizing antibody against Wuhan** | | |
| --- | --- | --- | --- | --- | --- | --- | --- | --- | --- | --- | --- | --- | --- | --- |
|  |  |  |  | No. | GMT, AU/mL | GCV, % |  | No. | GMT, U/mL | GCV, % |  | No. | GMT, NT_50_ | GCV, % |
| **Previous study 1** | 2-dose | Infection-naïve | 62 (40–69) | 85 | 6007 | 69.0 |  | 85 | 1200 | 62.2 |  | 85 | 417 | 69.3 |
| **Previous study 2** | 3-dose | Infection-naïve | 10 (9–11) | 43 | 22985 | 55.6 |  | 43 | 20421 | 32.6 |  | 43 | 672 | 89.7 |
| **Present study**  **(Total samples)** | 3-dose | Infection-naïve & previous infection | 173 (153–184) | 486 | 6156 | **132.6** |  | 486 | 6460 | **117.7** |  | 100 | 1164 | **232.7** |
| **Present study**  **(infection-naïve samples)** | 3-dose | Infection-naïve | 173 (153–184) | 429 | 5129 | **103.9** |  | 429 | 5500 | **93.8** |  | 82 | 843 | **158.1** |

Abbreviations: AU, arbitrary units; GCV, geometric coefficient of variation; GMT, geometric mean titer; NT_50_, 50% neutralization titer; IQR, interquartile range.

GCV (%) was calculated using the formula: $\surd(exp(variance)-1)\times100$.

Neutralizing antibody titers were tested against live viruses using HeLahACE2-TMPRSS2 cells in the present study and VeroE6TMPRSS2 cells in two previous studies.

Previous study 1: <https://dx.doi.org/10.1093/cid/ciab1048>

Previous study 2: <https://dx.doi.org/10.1016/j.ijsid.2023.01.023>
